# Outbreak of Chikungunya Fever in the Central Valley of Chiapas, Mexico

**DOI:** 10.1101/2024.10.09.24314897

**Authors:** Ana Luz Gonzalez-Perez, Ana Vazquez, Fernando de Ory, Anabel Negredo, Kenneth S. Plante, Jessica A. Plante, Pedro M. Palermo, Douglas Watts, Maria Paz Sanchez-Seco, Scott C. Weaver, Jose Guillermo Estrada-Franco

## Abstract

Chikungunya virus (CHIKV) was isolated from humans in an outbreak of a febrile illness during July and August 2015 in the central valleys of Chiapas, Mexico. Sera obtained from 80 patients were tested for CHIKV RNA by reverse transcriptase polymerase chain reaction (RT-PCR) and for IgM and IgG antibodies by an enzyme linked immunoassay and a commercial indirect immunofluorescence test for CHIKV and dengue virus (DENV). Of the 80 patients, 67 were positive, including 50 for RNA and 17 for IgM. In addition, one patient was coinfected with CHIKV-DENV and 40 patients were positive for IgG antibody to DENV. The clinical manifestations included a high fever, polyarthralgia, headache, myalgia, rash, digestive disorders, conjunctivitis, and adenopathy associated with cervical and axillary inguinal regions. Complete nucleotide sequences of two of the CHIKV isolates showed that they belonged to the Asian lineage but did not group with other Mexican CHIKV isolates from the Chiapas coast. Our findings documented that different Asian lineage strains of CHIKV were circulating simultaneously during the 2015 outbreak in the Central Valley of Chiapas, Mexico. The 2024 cases suggest an explosive scenario of re-emergence of thousands of new Chikungunya and dengue fever (DENF) cases associated with deaths, and a dangerous increase of the four DENV serotypes throughout the Americas, especially in South American countries that have shown a high influx of human migration to southern Mexico. In Mexico, the state of Chiapas and other southern regions are the most vulnerable.

**Author Summary:** The first ever recorded outbreak of Chikungunya virus (CHIKV) in the highlands (central valley of Chiapas) Mexico took place in 2015. Clinical data from 80 infected patients together with information provided from virus isolates offered a vision of the diseased and their response linked to different variables such as age, gender, association with pre-existing diseases of some patients, including CHIKF-DENF co-infection. The origins of the Tuxtla Gutierrez CHIKV viruses isolated in the study, including other Mexican isolates, were traced to viruses circulating in regions of Asia in Micronesia and Philippines. Also, we call the attention of the influx of migrants coming from countries having current explosive outbreaks of CHIKV and DENV in the Americas and into southern Mexico. Further we discuss vulnerabilities and point out specially to the geographic location of Chiapas State and its strategic position for attracting the human migrant population.

## Introduction

Chikungunya virus (CHIKV), *Togaviridae*, genus *Alphavirus*, is an arthropod-borne virus transmitted peri-domestically by *Aedes (Stegomyia) aegypti* and *Ae. (Stegomyia) albopictus* mosquitoes [1]. At least four genetic lineages of CHIKV have been described including West African, East/ Central/South African, Asian, and Indian Ocean [2].

The first documented outbreak of chikungunya fever (CHIKF) associated with autochthonous transmission in the Americas was reported during December of 2013 on the Caribbean Island of St. Martin [3]. In 2014, the Pan American Health Organization (PAHO) estimated more than a million cases of CHIKF in 50 countries in the Americas [4]. The disease manifestations include fever ˃38°C, rash and severe arthralgia that is disabling, and patients may also experience a chronic phase with continuing articular and periarticular inflammation lasting for months or years that is seldom fatal [5].

In 2004, CHIKV began to cause major outbreaks that affected humans in substantial portions of Africa, the Indian Ocean Basin, Asia, Europe, and more recently the Americas [6]. The spread of CHIKV was exacerbated by infected people travelling from regions of active virus transmission [7]. Millions were affected, and this scenario was further complicated in some areas, due in part to the adaptive mutations in the CHIKV viral genome that resulted in changes in the E1 (A226V) [8,9] and E2 (several substitutions) glycoproteins [10]. These mutations enhanced infection of *Ae. albopictus* mosquitoes, and an increase in the vector competence of this species for CHIKV. As a result, European outbreaks of CHIKF in Italy and France and in islands of the Indian Ocean [11] were associated with the distribution of *Ae. albopictus* mosquitoes, thus demonstrating the emerging importance of this species as a potential vector of CHIKV [12]. The ability of both *Ae. aegypti* and *Ae. albopictus* to serve as efficient vectors represents a highly efficient vectorial system for the transmission of CHIKV worldwide [13].

In Mexico, the first traveler case of CHIKF was reported in Jalisco state in May 2014 [14], and the first autochthonous case was diagnosed in Coastal Chiapas, Mexico, in November 2014 [15]. In a period of 10 years (2014 – 2024), Mexicós Health Ministry recorded 776 cases of CHIKF in the State of Chiapas [16]. According to the Pan American Health Organization (PAHO), 273,685 CHIKF cases were recorded in 2022 and 410,754 CHIKF cases were recorded during 2023 in 17 countries of the Americas, including 419 deaths, representing a 43% increase in the number of cases. In 2024, 186,274 cases with 60 deaths had been recorded by the epidemiological week 14 [17]. Clearly there is an escalation of CHIKV cases reported mainly in South American countries with a high migratory trend in the south Chiapas Mexican border [18].

The occurrence during early 2015 of unusual dengue-like cases in the medical community of the Metropolitan region of Tuxtla Gutierrez, the capital State of Chiapas, suggested the outbreak represented an extension of the first epidemic of Mexican CHIKV recorded in the Statés coastal region [19]. Here we describe the epidemiology, clinical, and molecular observations of the first major epidemic of CHIKF that occurred during the summer of 2015 in the central valleys of Chiapas, Mexico. Phylogenetic studies indicated that the outbreaks in Mexico were caused by strains of the CHIKV Asian lineage [20], and *Ae. aegypti* was incriminated as the principal vector [21].

### Study area: The Central Valley of Chiapas

The study was carried out in the Central Valley of Chiapas during July and August of 2015 at three sites, two health care facilities: 1) at the General Hospital “Dr. Belisario Domínguez”, Institute of Security and Social Services of State Workers ISSSTE, located in Tuxtla Gutiérrez (16 ° 45’11 “N 93 ° 06’56” W; altitude 522 meters), and including patients from the municipalities of Berriozabal, Chiapa de Corzo, and Suchiapa (Fig 1 and Fig 2) the Rural Hospital of Solidarity Opportunities, Mexican Institute of Social Security IMSS Number 31, located in Ocozocoautla de Espinoza (16 ° 45’00 “N 93 ° 22’00” W; 818 altitude meters) at 18.7 km west of Tuxtla Gutierrez. The third site was Miguel Álvarez del Toro Zoo (ZooMAT) located in Tuxtla Gutierrez.

**Fig 1.**
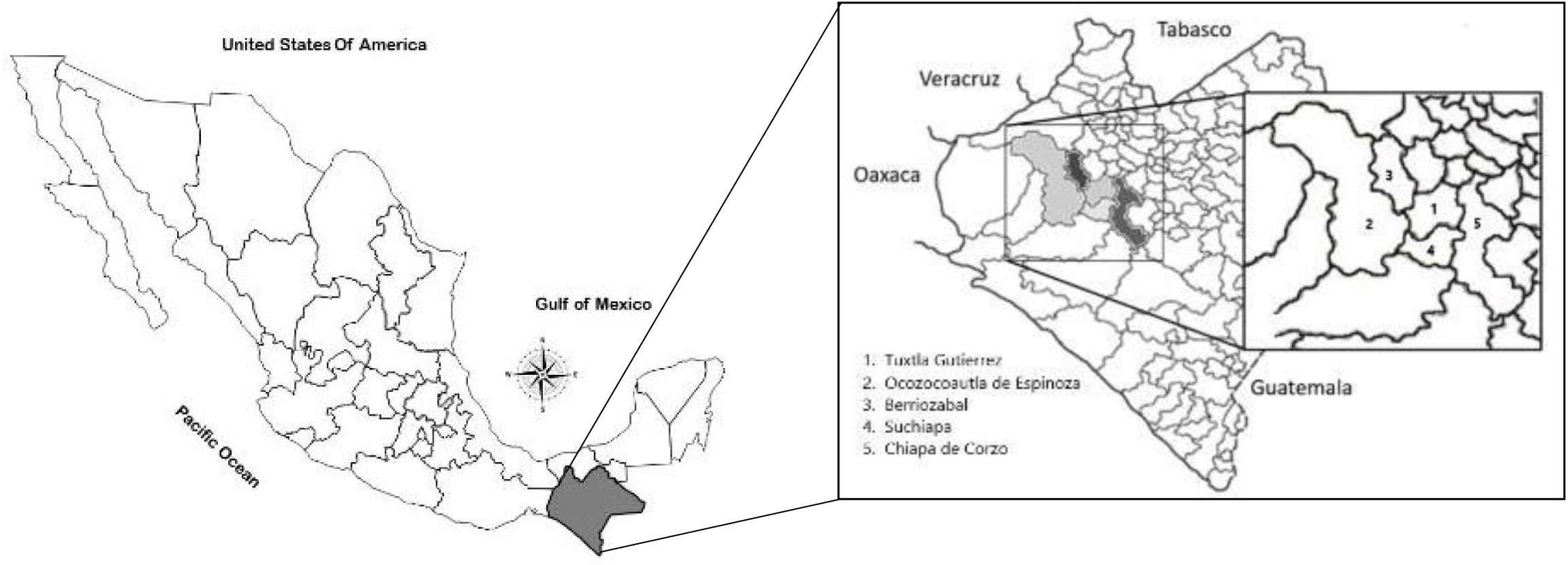
Map of Chiapas, Mexico showing the sites where serum samples were obtained to test for Chikungunya virus in 2015.

**Fig 2.**
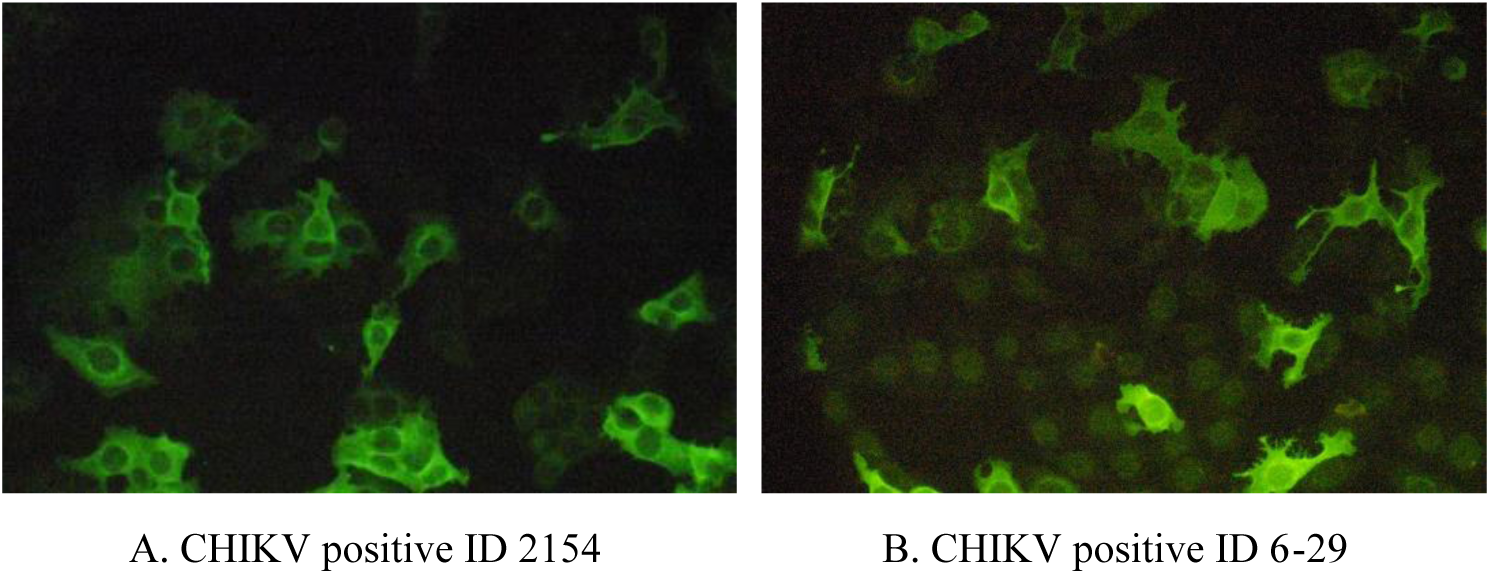
The results for testing sera sample obtained from suspected chikungunya fever cases by a commercial indirect immunofluorescent antibody assay during an outbreak of chikungunya fever in 2015 in Chiapas, Mexico.

The climate in the Central Valley is warm and humid, with an average annual temperature of 25.4°C and annual rainfall of 1,200 mm [22].

### Patient Selection Criteria

Blood samples were collected from patients who presented at the health care facilities and samples from suspected zoo workers, using a case definition of suspected CHIKF including a fever ˃38°C and arthritis/arthralgia with an acute onset that was not explained by other medical conditions, and residing in or visiting the epidemic areas during two weeks prior to the onset of illness [23].

### Patient Data Collection

A questionnaire was designed to record the following variables: a) Epidemiological and demographic: age, sex, contact with relatives, neighbors, friends at school and/or work contacts, previous days of illness, and recent travel; b) Clinical: detailed description of the symptoms and signs; c) Laboratory: blood biometry, serology, and RT-PCR results; d) Environmental: community environment characteristics.

### Sample Collection

Venipuncture blood samples were collected and centrifuged at 1207 x g to obtain serum that was aliquoted and stored at -80°C. Samples were sent for diagnostic testing and nucleotide sequencing to the World Reference Center for Emerging Viruses and Arboviruses at the University of Texas Medical Branch (UTMB), Galveston, Texas, USA; to the National Center for Microbiology (CNM), Instituto de Salud Carlos III (ISCIII), Majadahonda, Spain and to the Virology Laboratory at the University of Texas at El Paso (UTEP), Texas.

### Blood Biometry

Hematological values were determined using a hematology analyzer Advia® 120 (Siemens Healthcare Diagnostics, Erlangen, Germany).

### Molecular assays

They were conducted through two protocols: 1) Viral RNA was extracted from sera using the ZR-96 Viral RNA Kit (Zymo Research, Orange, CA, USA) according to the manufacturer’s protocol. A one-step quantitative reverse transcription PCR (qRT-PCR) was performed by using the TaqMan RNA-to-Ct 1-step Kit (Applied Biosystems, San Francisco, CA, USA) [24]. 2) To extend the screening, RNA was extracted from the sera using the QIAamp viral RNA kit (Qiagen, Valencia, CA) following the manufacturer’s protocol. Next, RT-PCR targeting the alphavirus nsP4 gene was used to detect CHIKV RNA, as previously described [25]. The results were confirmed with a specific nested PCR for CHIKV, which was directed to the E1 gene [26]. Subsequently, all samples were screened by qRT-PCR for CHIKV (Realstar CHIKV RT-PCR kit, Altona Diagnostic, Hamburg, Germany). All negative samples were also tested for other etiologic agents, including bunyaviruses, arenaviruses, hantaviruses, and phleboviruses using in-house RT-PCR techniques [27–29].

## Methods

### Ethical considerations

The Bioethics Committee of the Faculty of Medicine of the Autonomous University of the State of Mexico (Approval No. 011/2015) approved the study. Written informed consent was obtained from all patients who enrolled in this study. Minor patients participated with the prior authorization of their parents. Patients’ personal information was anonymized before analyzing patient data, including demographic characteristics, clinical signs and symptoms.

### Serology assays

All samples were tested for CHIKV antibody using enzyme-linked immunosorbent assay (ELISA) [30]. An IgM antibody-capture (ELISA) was performed using a chimeric Eilat-CHIK containing the nonstructural protein genes of Eilat virus and structural protein genes of CHIKV, which produces virions indistinguishable structurally from those of CHIKV [31]. Additionally, sera were screened by a commercial indirect immunofluorescent (IIF) anti-Chikungunya Virus IIFT IgG and IgM (Euroimmun AG, Lübeck, Germany) [32, 33] and the IIF assay Arboviral Fever Mosaic IgG and IgM, Euroimmun, Germany [34, 35]. Also, the samples were tested to determine antibody titers by a dengue IgM capture ELISA, Panbio® and dengue indirect IgG ELISA, Panbio®, Queensland, Australia [36].

### Sequencing and phylogenetic analysis

Two CHIKV positive sera (ID 2107 and ID 2115) were selected for Illumina® Bio Systems nucleotide sequencing. Available CHIKV sequences encoding both open reading frames of the genome were retrieved from the Genbank database. CHIKV sequences without published references were excluded. A total of 143 representative strains were selected for the phylogenetic analysis including our CHIKV strains 2107 and 2115. Sequences were trimmed to remove the 5’ and 3’ untranslated genome regions (UTRs) and were aligned using Clustal Omega in MegAlign Pro version 12.0.0 (DNASTAR Inc, Madison, WI). Maximum likelihood analysis was performed using the phangorn package [37] in R version 3.4.2 [38]. The GTR+G+I model of nucleotide substitution was selected from the 24 options analyzed as part of the model test analysis in phangorn based on minimizing the AIC score. Bootstrapping was performed with 1,000 iterations. The resulting tree was visualized using iTOL version 4.3.3 [39].

## Results

Among the sera obtained from 80 patients, 46 were collected in Tuxtla Gutierrez, 31 in Ocozocoautla de Espinoza and 3 from the municipalities of Chiapa de Corzo, Berriozabal and Suchiapa (Fig 1). Of the 46 samples collected in Tuxtla Gutierrez, 7 were from ZooMAT workers. We conducted two assay protocols on the 80 samples at UTMB and AEVI-CNM-ISCIII and obtained 50 positives for CHIKV RNA RT-PCR and 17 samples were positive by ELISA for CHIKV IgM antibody. The 17 IgM positive samples were reconfirmed by IIF for Chikungunya IgM antibody. Of the seven samples from patients related to the ZooMAT, three were positive for CHIKV RNA by RT-PCR, qRT-PCR and three were positives CHIKV IgM or IgG antibody by ELISA.

The assay of the 80 serum samples by RT-PCR for arenavirus, hantavirus, bunyavirus and phlebovirus viral RNA was negative. One CHIKV RNA positive sample and one CHIKV RNA negative were positive for dengue IgM antibody by ELISA. Also, the results revealed that 40 of the 80 patients had DENV IgG antibody as evidence of a previous infection.

### Viral isolation

Seventeen of the 50 CHIKV PCR-positive samples were tested for virus isolation in C6/36 cells. CHIKV was isolated from all seventeen samples based on the detection of cytopathic effects on day 6 post-inoculation and were also positive for CHIKV by IFA as more conclusive results.

### Sequencing and Phylogenetic Analysis

The genomes of two CHIKV isolates, ID 2107 and ID 2115, were fully sequenced and the assembled complete genome sequences were submitted to GenBank under the accession numbers 1036 and 1037. A phylogenetic analysis was performed, along with 143 geographically and temporally diverse CHIKV sequences from GenBank encompassing Asian, East/Central/South African (ECSA), Indian Ocean and West African lineage strains of CHIKV (S1 Table). A maximum-likelihood tree was generated using complete open reading frame sequences with the 5’ and 3’ UTRs removed. The results showed that both isolates (ID 2107 and ID 2115) clustered with the Asian lineage and with the other CHIKV isolates obtained from the Americas and the Caribbean (Fig 3). The uncorrected pairwise distances between both 2107 and 2115 and the other Asian lineage isolates from the Americas and Caribbean ranged from 0.0003 (0.03% nucleotide sequence divergence) to 0.0016. Their distance to the other Asian lineage strains varied from 0.0015 for strains thought to immediately precede the introduction of CHIKV to the Caribbean, such as the Yap 2013 isolate, and up to 0.0279 for the 1973 Indian isolate. Their distances from the ECSA strains ranged from 0.0559 to 0.0654, and their distances from all including West African strains ranged from 0.1565 to 0.1583.

**Fig 3.**
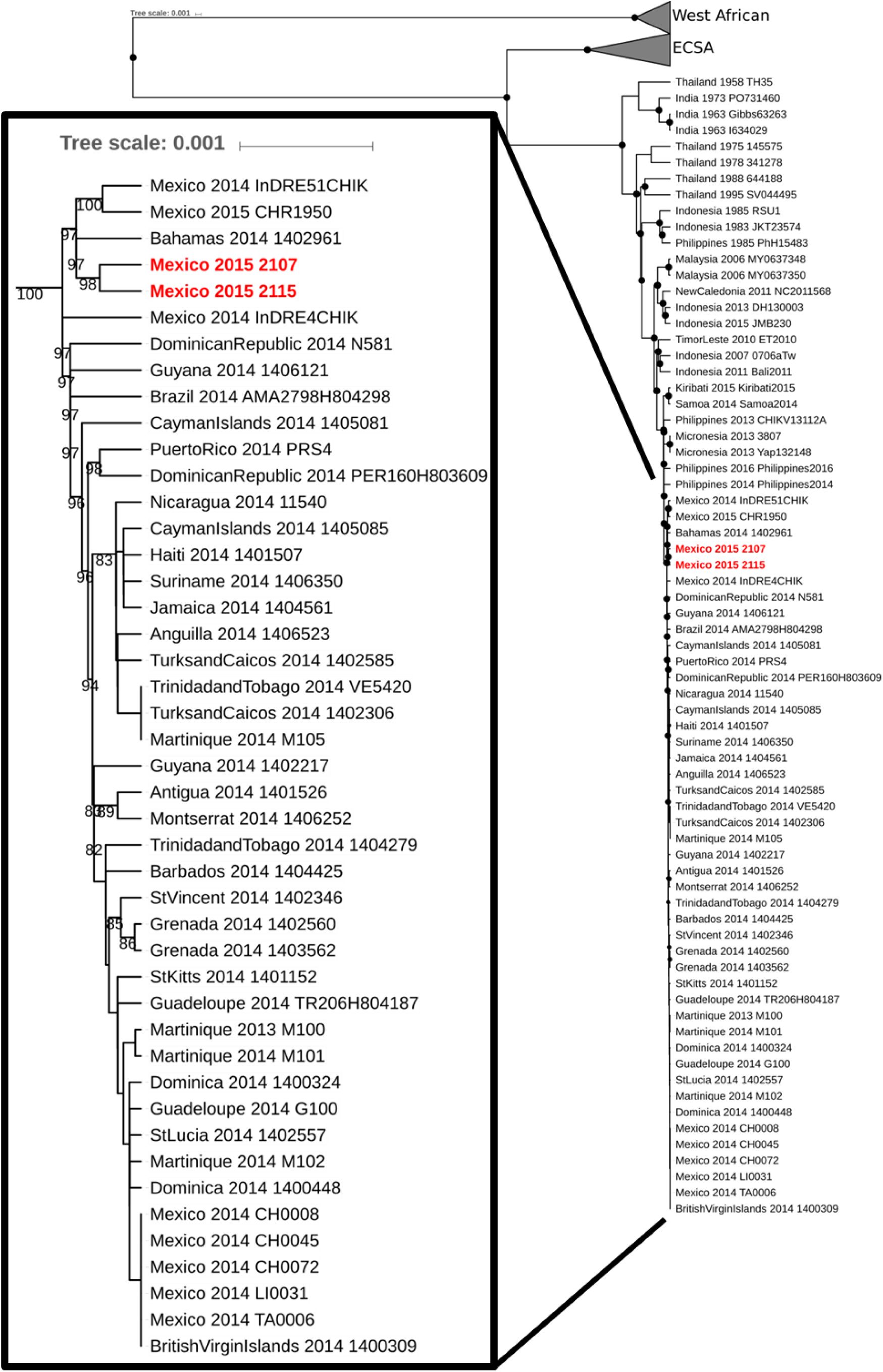
Phylogenetic analysis of CHIKV isolates obtained from patients during an outbreak of chikungunya fever in Chiapas, Mexico. Maximum-likelihood tree based on the full genome sequences (minus the 5’ and 3’ UTRs) of 143 geographically and temporally diverse strains of CHIKV from GenBank, along with strains 2107 and 2115 (highlighted with red and bold text). Branch lengths are scaled relative to substitutions. Bootstrapping was performed with 1,000 iterations; values ≥80 are represented as closed black circles at the node in the full phylogenetic tree (right), and are listed as text in the expanded view of the Asian lineage strains from the Americas and Caribbean (left).

There were 110 variable nucleotide positions among the Asian lineage CHIKV sequences from the Americas. The 2107 and 2115 strains differed from the consensus sequence at 11 positions (five in 2107, two in 2115, and four mutual positions for both strains 2107 and 2115) (Table 1). Only one of these changes encoded an amino acid substitution in nsP3 (H377P) which was in the hypervariable region and was unique to strain 2115 (Table 1). It should be noted, however, that all of the West African lineage isolates analyzed possess a leucine at this position and five of the 26 Asian lineages isolates from Asia did not contain a deletion in this codon 377. Neither strain 2107 nor 2115 encoded the *Ae. albopictus*-adaptive E1 [9] or E2 [11] substitutions identified in some of the Indian Ocean Lineage CHIKV strains.

**Table 1.**
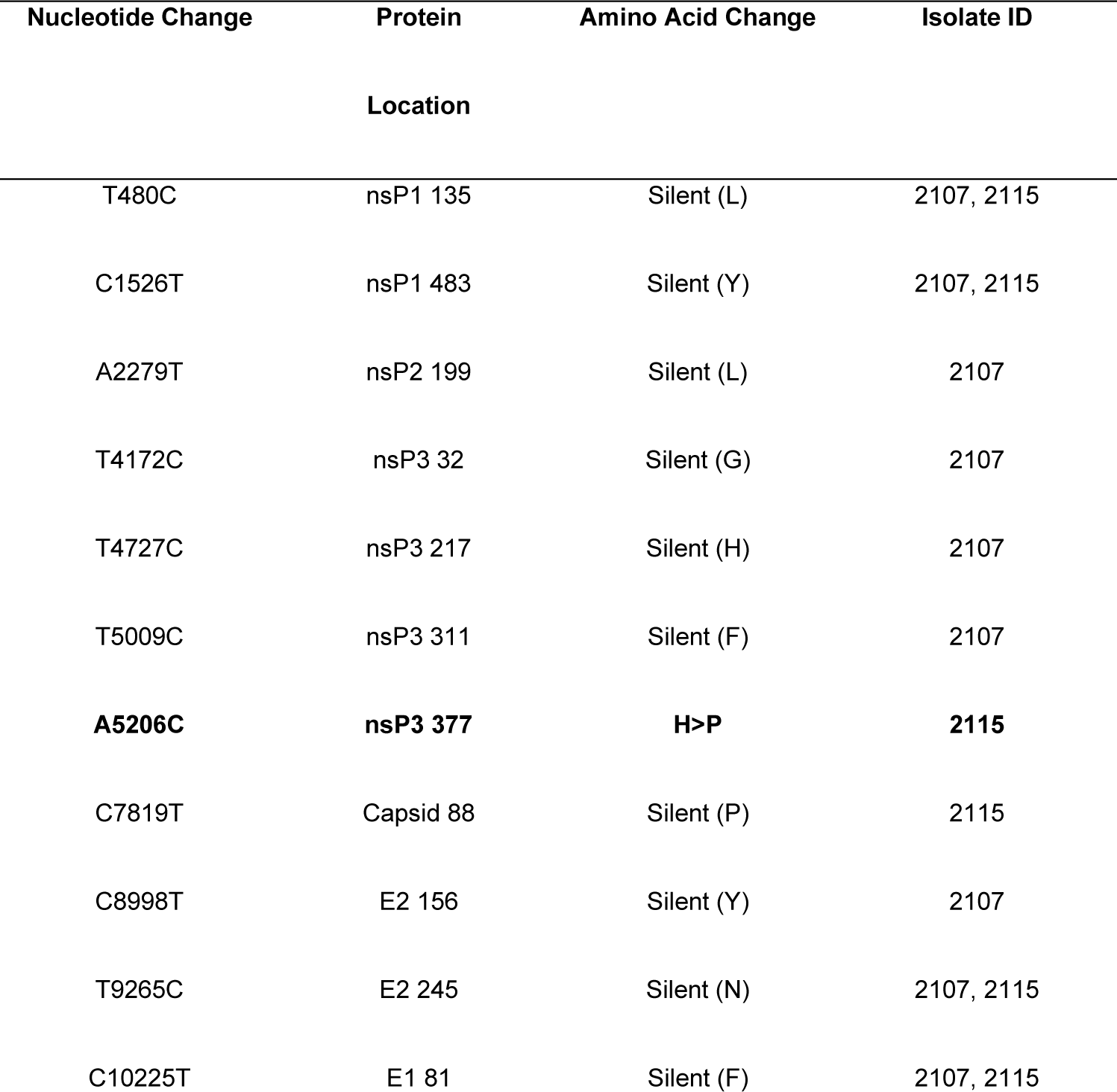
Mutations in complete chikungunya virus ORF nucleotide sequences for chikungunya virus isolates (ID 2017 and ID 2115) from Chiapas patients during an outbreak 2015 in Chiapas, Mexico. Summary of nucleotide-level variation between chikungunya virus isolates 2107 and 2115 and 43 other New World Asian lineages strains of chikungunya virus. Nucleotide and amino acid positions are based on Accession KP164567.1

Amino acid variation was also analyzed among CHIKV isolates ID 2107 and ID 2115, and the other eight Mexican CHIKV isolates from other studies with complete coding sequences available in GenBank (Table 2). Other than the nsP3 H377P change detected in 2115, only one amino acid in nsP3 varied among Mexican isolates: L340S, detected only in CH-R-1950.

**Table 2.** Amino acid variation among Mexican chikungunya virus (CHIKV) isolates obtained during an outbreak of chikungunya fever in 2015 in Chiapas, Mexico. Summary of amino acid variation between all Mexican isolates of CHIKV with full coding sequences deposited in Gene Bank, in addition to 2107 and 2115. Consensus determined by alignment of 45 New World Asian lineage strains of CHIKV. Bold indicates non-consensus reside. Amino acid positions are based on Accession KP164567.1.

| Protein name | AA position | Strains |  |  |  |  |  |  |  |  |  |
| --- | --- | --- | --- | --- | --- | --- | --- | --- | --- | --- | --- |
|  |  | 2107 | 2115 | CH0008 | CH0045 | CH0072 | InDRE 4CHIK | InDRE 51CHIK | LI0031 | TA0006 | CH-R-1950 |
| nsP3 | 340 | Leu | Leu | Leu | Leu | Leu | Leu | Leu | Leu | Leu | **Ser** |
| nsP3 | 377 | His | **Pro** | His | His | His | His | His | His | His | His |
| nsP4 | 274 | Asn | Asn | Asn | Asn | Asn | **Asp** | Asn | Asn | Asn | Asn |
| nsP4 | 399 | Ser | Ser | Ser | Ser | Ser | Ser | **Cys** | Ser | Ser | Ser |
| nsP4 | 432 | Met | Met | Met | Met | Met | **Thr** | Met | Met | Met | Met |
| E3 | 42 | Pro | Pro | Pro | Pro | Pro | Pro | Pro | Pro | Pro | **Ser** |
| E2 | 113 | Val | Val | **Ala** | **Ala** | **Ala** | Val | Val | **Ala** | **Ala** | Val |

Interestingly, although the Mexican isolates were evenly split between valine and alanine at E3-113, it is worth noting that all other strains considered in the phylogeny possessed a valine at this position except for the 2014 isolate from the British Virgin Islands isolate, which contains an alanine.

### Blood biometrics

Blood count abnormalities were found among the CHIKV positive patients. Samples had changes in one or all three cell types (leukopenia, lymphopenia and neutropenia) that help to differentiate CHIKF, especially in the acute phase [40] (Table 3).

**Tabla 3.** Hematological abnormalities in 67 chikungunya fever patients during an outbreak of chikungunya fever in 2015 in Chiapas, Mexico.

| Blood formula | % (n=67) |
| --- | --- |
| Leukopenia (<4.50 miles/μl) | 13 (9) |
| Leukocytosis (>10.50 miles/μl) | 9 (6) |
| Erythrocytosis (>5.0 million/μl) | 67 (11) |
| Hemoglobin Low (<12.5 g/dl) | 13 (9) |
| Hematocrit Low (<37.0 %) | 2 (1) |
| Lymphopenia (<24 miles/UI) | 52 (35) |
| Lymphocytosis (>38 miles/UI) | 12 (8) |
| Monocytopenia (<4.0 miles/UI) | 68 (11) |
| Monocytosis (>10.0 miles/UI) | 8.5 (6) |
| Thrombocytopenia (<50.0 miles/μl) | 7.4 (5) |

### Epidemiological variables

#### Demographic

Of the 67 CHIK-positive patients, 40% were male and 60% were female. Most of the cases of CHIKF occurred among the 10-59 years old group (Table 4). Among occupations, 34 (51%) were workers (professional, employee, crafts), 19 (28%) students, 11 (16%) housemakers, two (3%) retired, and one (2%) was a preschooler patient.

**Table 4.** Incidence of chikungunya virus (CHIKV) infection by age and gender during a chikungunya fever outbreak during 2015 in Chiapas, Mexico.

| Age<br>(Years) | Infected CHIKV<br>Female (%) | Infected CHIKV<br>Male (%) | Total Infected<br>CHIKV% |
| --- | --- | --- | --- |
| 1-9 | 4 (67) | 2 (3) | 6 (100) |
| 10-19 | 10 (53) | 5 (26) | 15 (79) |
| 20-29 | 5 (41) | 4 (33) | 9 (75) |
| 30-39 | 6 (50) | 3 (25) | 9 (75) |
| 40-49 | 10 (63) | 5 (31) | 15 (94) |
| 50-59 | 6 (55) | 4 (36) | 10 (91) |
| 60-69 | 1 (100) | 0 (N/A) | 1 (100) |
| 70-79 | 0 | 0 (N/A) | 0 (0) |
| 80-89 | 0 | 2 (67) | 2 (67) |
| Total | 42 | 25 | 67 |

#### Clinical-epidemiological findings

During the initial phase, 40% of CHIKV-positive patients were misdiagnosed by the primary care medical staff. The treating health personnel recorded signs and symptoms due to other etiologies, including 60% who were diagnosed with DENF, 25% with other types of illnesses such as respiratory diseases (15%), stress and chronic exhaustion (5%), common headache (3%), food allergies (2%); and only 15% were properly diagnosed as CHIKF cases. CHIKV positive patientśsigns/symptoms included fever 38 °C - 41.0 °C (x̅= 38.3°C) with 2-5 days of evolution. The dominant signs/symptoms observed in the patients were polyarthralgia in 67 people (83.3%; 95% CI 75%-91%) of 1-5 days duration, headache 53 patients (66%; 95 CI 55%-76%) evolving within a range of 1-7 days, myalgia 46 patients (57.4%; 95% CI 46%-68%) in 2-3 days, conjunctivitis 42 individuals (52.4%; 95% CI 41%-63%) progressing in 1-4 days, and exanthema 42 patients (52.4%; 95% CI 41%-63%) unfolding in 1-6 days (Table 5). Symmetric bilateral arthralgia was present in 52 individuals (64.8%; 95% CI 54%-75%) and a severe degree of pain was observed in 36 individuals (45%;95% CI 34%-55%). Nine (11.7%; 95% CI 5.2%-18%) patients had pain in all joints of the body. In 33 persons affected joints involved the upper (41.3%;95% CI 30%-52%) (shoulder, elbow, wrist, hand, fingers) and in 23 (29%; 95% CI 19%-38%) patients lower limbs (hip, knee, ankle. foot, toes). Acute cutaneous manifestations were present in 42 patients (52.4%; 95% CI 41%-63%), including a rash that presented as maculopapular and pruritic, pruritic, maculopapular and diffuse (Table 6). Bullous and pruritic rash was observed in a school-age patient. The rash developed mainly in arms in 30 persons (37.6%; 95% CI 27%-48%), 16 patients in legs (20.3%; 95% 11%-29%), and in 15 persons (19.1%; 95% CI 10%-27%) (Table 7). Five patients reported having burning sensation in the skin, three of them in the soles and two in the palms (Table 7).

**Table 5.** Clinical manifestation of the patients infected during a chikungunya fever outbreak of 2015 in Chiapas, Mexico.

| Signs/<br>Symptoms | % Female | (n=67) | 95% CI | % Male | %(n=67) | 95% CI | % Total<br>(n=80) | 95% CI |
| --- | --- | --- | --- | --- | --- | --- | --- | --- |
| Arthralgias | 62.7 (42) | 52.4 (42) | 41%-63% | 37.3 (25) | 31.4 (25) | 21%-41% | 83.7 (67) | 75%-91% |
| Headache | 60.4 (32) | 40.1 (32) | 29%-50% | 39.6 (21) | 26.5 (21) | 17%-36% | 66 (53) | 55%-76% |
| Myalgia | 56.2 (26) | 32.7 (26) | 22%-42% | 43.5 (20) | 25.3 (20) | 16%-34% | 57.4 (46) | 46%-68% |
| Conjunctivitis | 59.5 (25) | 31.4 (25) | 21%-41% | 40.5 (17) | 21.6 (17) | 12%-30% | 52.4 (42) | 41%-63% |
| Exanthema | 64.3 (27) | 33.9 (27) | 23%-44% | 35.7 (15) | 19.1 (15) | 10%-27% | 52.4 (42) | 41%-63% |
| Depression | 63.4 (26) | 32.7 (26) | 22%-42% | 36.6 (15) | 19.1 (15) | 10%-27% | 51.2 (41) | 40%-62% |
| Abdominal pain | 69.5(25) | 31.4 (25) | 21%-41% | 30.5 (11) | 14.1 (11) | 7%-21% | 45 (36) | 34%-55% |
| Retro eye pain | 57.6 (19) | 24 (19) | 15%-33% | 42.4 (14) | 17.9 (14) | 9.9%-26% | 41.3 (33) | 30%-52% |
| Adenopathy | 71.4 (20) | 25.3 (20) | 16%-34% | 28.6 (8) | 10.4 (8) | 4.3%-17% | 35.1 (28) | 24%-45% |
| Photophobia | 50.0 (14) | 17.9 (14) | 10%-25% | 50.0 (14) | 17.9 (14) | 9.9%-26% | 35.1 (28) | 24%-45% |
| Nausea | 70.4 (19) | 24 (19) | 15%-33% | 29.6 (8) | 10.4 (8) | 4.3%-17% | 33.9% (27) | 23%-44% |
| Tonsillitis | 66.7 (18) | 22.8 (18) | 14%-32% | 33.3 (9) | 11.7 (9) | 5.2%-18% | 33.9% (27) | 23%-44% |
| Paresia | 58.3 (14) | 17.9 (14) | 10%-26% | 41.7 (10) | 12.9 (10) | 6.1%-20% | 30.2 (24) | 20%-40% |
| Cough | 50.0 (11) | 14.1 (11) | 7%-21% | 50.0 (11) | 14.1 (11) | 7%-21% | 27.7 (22) | 18%-37% |
| Diarrhea | 68.2 (15) | 19.1 (15) | 10%-27% | 31.8 (7) | 9 (7) | 0.3%-15% | 27.7 (22) | 18%-37% |
| Nasal<br>congestion | 66.7 (14) | 17.9 (14) | 10%-26% | 33.3 (7) | 9 (7) | 0.3%-15% | 26.5 (21) | 17%-36% |
| Dyspnoea | 60.0 (12) | 15.4 (12) | 8%-23% | 40.0 (8) | 10.4 (8) | 4.3%-17% | 25.3 (20) | 16%-34% |
| Otitis | 64.7 (11) | 14.1 (11) | 7%-21% | 35.3 (6) | 8 (6) | 2.6%-13% | 21.6 (17) | 12%-30% |
| Facial<br>hyperemia | 82.4 (14) | 17.9 (14) | 9.9%-26% | 17.6 (3) | 4.3 (3) | 0.6%-8.7% | 21.6 (17) | 12%-30% |
| Petechiae | 62.5 (10) | 12.9 (10) | 6.1%-20% | 37.5 (6) | 8 (6) | 2.6%-13% | 20.3 (16) | 11%-29% |
| Inappetence | 73.3 (11) | 14.1 (11) | 7%-21.0% | 26.7 (4) | 5.5 (4) | 1.2%-10% | 19.1 (15) | 10%-27% |
| Abnormal muscle sensitivity | 53.3 (8) | 10.4 (8) | 4.3%-17% | 46.7 (7) | 9 (7) | 0.3%-15% | 19.1 (15) | 10%-27% |
| Pharyngitis | 78.6 (11) | 14.1 (11) | 7.0%-21.0% | 21.4 (3) | 4.3 (3) | 0.6%-8.7% | 17.9 (14) | 9.9%-26% |
| Puke | 75.0 (9) | 11.7 (9) | 5.2%-18% | 25 (3) | 4.3 (3) | 0.6%-8.7% | 15.4 (12) | 7.9%-23% |
| Ulcers | 41.7 (5) | 7 (5) | 1.9%-12% | 58.3 (7) | 9 (7) | 0.3%-15% | 15.4 (12) | 7.9%-23% |
| Agitation | 72.7 (8) | 10.4 (8) | 4.3%-17% | 27.3 (3) | 4.3 (3) | 0.6%-8.7% | 14.1 (11) | 7.0%-21% |
| Dizziness | 60.0 (6) | 8 (6) | 2.6%-13% | 40 (4) | 5.5 (4) | 1.2%-10% | 12.9 (10) | 6.1%-20% |
| Swelling of limbs | 55.5 (5) | 6.7 (5) | 1.9%-12% | 44.6 (4) | 5.5 (4) | 1.2%-10% | 11.7 (9) | 5.2%-18% |
| Drowsiness | 66.7 (6) | 8 (6) | 2.6%-13% | 33.3 (3) | 4.3 (3) | 0.6%-8.7% | 11.7 (9) | 5.2%-18% |
| Gum pain | 50 (4) | 5.5 (4) | 1.2%-10% | 50 (4) | 5.5 (4) | 1.2%-10% | 10.4 (8) | 4.3%-17% |
| Tremors | 62.5 (5) | 6.7 (5) | 1.9%-12% | 37.5 (3) | 4.3 (3) | 0.6%-8.7% | 10.4 (8) | 4.3%-17% |
| Burning in skin | 80.0 (4) | 5.5 (4) | 1.2%-10% | 20 (1) | 1.8 (1) | 0.2%-4.7% | 6.7 (5) | 1.9%-12% |
| Bleeding | 75 (3) | 4.3 (3) | 0.6%-8.7% | 25 (1) | 1.8 (1) | 0.2%-4.7% | 5.5 (4) | 1.2%-10% |
| Jaundice | 100 (2) | 3 (2) | 0.1%-6.0% | N/A(0) | 0 (0) | 0.2% | 3 (2) | 0.1%-6.0% |
| Confusion | 100 (2) | 3 (2) | 0.1%-6.0% | N/A(0) | 0 (0) | 0.2% | 3 (2) | 0.1%-6.0% |

CI 95 % Confidence interval

**Table 6.** Characteristics of the arthralgia for chikungunya fever cases during an outbreak of in 2015 in Chiapas, Mexico.

| Characteristic arthralgia | % (n=67) | % (n=80) | 95% CI |
| --- | --- | --- | --- |
| **Distribution** |  |  |  |
| Arthralgia bilateral | 77.6 (52) | 64.8 (52) | 54%-75% |
| Arthralgia unilateral | 22.4 (15) | 19.1 (15) | 10%-27% |
| Arthralgia symmetrical | 77.6 (52) | 64.8 (52) | 54%-75% |
| **Degree of pain** |  |  |  |
| Mild Pain | 5.9 (4) | 5.5 (4) | 1.2%-10% |
| Moderate Pain | 40.3 (27) | 33.9 (27) | 23%-44% |
| Severe Pain | 53.7 (36) | 45 (36) | 34%-55% |

| Anatomical sites joint |  |  |  |
| --- | --- | --- | --- |
| Arms | 49.2 (33) | 41.3 (33) | 30%-52% |
| Shoulder | 6 (4) | 5.5 (4) | 1.2%-10% |
| Elbow | 8.9 (6) | 8.0 (6) | 2.6%13% |
| Wrist | 32.8 (22) | 27.7 (22) | 18%-37% |
| Hand | 22.3 (15) | 19.1 (15) | 10%-27% |
| Fingers | 14.9 (10) | 12.9 (10) | 6.1%-20% |
| Legs | 34.3 (23) | 29 (23) | 19%-38% |
| Hips | 25.3 (7) | 9% (7) | 0.3%-15% |
| Knee | 23.8 (16) | 20.3% (16) | 11%-29% |
| Ankle | 23.8 (16) | 20.3% (16) | 11%-29% |
| Foot | 23.8 (16) | 20.3% (16) | 11%-29% |
| Jaw | 8.9 (6) | 8.0% (6) | 2.6%1.3% |
| Neck | 8.9 (6) | 8.0% (6) | 2.6%1.3% |
| Chest | 7.4 (5) | 6.7% (5) | 1.9%-12% |
| Trunk | 25.3 (17) | 21.6% (17) | 12%-30% |
| Spine | 4.4 (3) | 4.3% (3) | 0.6%-8.7% |
| Waist | 10.4 (7) | 9 (7) | 0.3%-15% |
| All Joints | 13.4 (9) | 11.7% (9) | 5.2%-18% |

CI 95 % Confidence interval

**Table 7.** Characteristics of the exanthema in 42 patients of the 67 positive chikungunya fever cases during an outbreak of in 2015 in Chiapas, Mexico.

| Characteristic exanthema | % (n=42) | % (n=80) | 95% CI |
| --- | --- | --- | --- |
| **Distribution** |  |  |  |
| Maculopapular | 7.14 (3) | 4.3 (3) | 0.6%-8.7% |
| Maculopapular and Pruritic | 23.8 (10) | 12.9 (10) | 6.1%-20% |
| Pruritic | 21.4 (9) | 11.7 (9) | 5.2%-18% |
| Bullosa and Pruritic | 2.4 (1) | 1.8 (1) | 0.2%-4.7% |
| Difuse | 7.1 (3) | 4.3% (3) | 0.6%-8.7% |
| Difuse and Pruritic | 2.4 (1) | 1.8% (1) | 0.2%-4.7% |
| **Anatomical sites exanthema** |  |  |  |
| Face | 11.9 (5) | 6.7 (5) | 1.9%-12% |
| Neck | 2.3 (1) | 1.8 (1) | 0.2%-4.7% |
| Trunk | 33.3 (14) | 17.9 (14) | 9.9%-26% |
| Arms | 71.4 (30) | 37.6 (30) | 27%-48% |
| Hand | 7.1 (3) | 4.3 (3) | 0.6%-8.7% |
| Chest | 11.9 (5) | 6.7% (5) | 1.9%-12% |
| Abdomen | 35.7 (15) | 19.1% (15) | 10%-27% |
| Back | 19 (8) | 10.4% (8) | 4.3%-17% |
| Legs | 38.1 (16) | 20.3% (16) | 11%-29% |
| Feet and Soles | 4.7 (2) | 3% (2) | 0.1%-6% |
| Entire Body | 2.3 (1) | 1.8% (1) | 0.2%-4.7% |

CI 95 % Confidence interval

Adenopathy was observed in 28 CHIKF patients (35.1%; 95% CI 24%-45%) 41.8%, including cervical (retroauricular, neck), axillary and inguinal region. Twelve patients involved a single anatomical site and 16 in two or more anatomical areas. Four CHIKF patients required hospitalization and 14 persons reported comorbidities, including three patients with diabetes, five with hypertension, two persons with diabetes and hypertension, one with muscular dystrophy, one patient disc herniation and cystitis, one patient with Horner syndrome and, one with hypertension and one with vertigo.

#### Environment characteristics

Seventy four percent CHIKF cases lived in urban areas and 26% lived in suburban-rural areas. Fifty-one percent had contact with CHIKF like homes, work, school or in the same locality. Fifty-four percent had periodic contact with bodies of water. The characteristics of the water for 40% of the cases was sewage, 30% clean running water (streams, lakes) and 6.6% standing water. Twenty two positive patients lived, worked or studied near a dump or cemetery. Fifty three patients lived near areas with abundant vegetation. All of these premises were located at distance of ≤150 meters of radius. At the ZooMAT we obtained six positive samples wild forest reserve area with a large population of vertebrate wild fauna and many invertebrates particularly *Aedes albopictus* and *Ae. aegypti* mosquitoes in abundance within the reserve.

### Discussion

We report the first CHIKV isolates from febrile patients in the Central Valley of Chiapas State, Mexico, including five municipalities: Tuxtla Gutiérrez, Ocozocoautla de Espinoza, Berriozabal, Chiapa de Corzo, and Suchiapa. Isolates were obtained from patients who resided about 130 km away from the first Mexican autochthonous recorded CHIKF case in 2014 [16].

Phylogenetic analysis showed that our CHIKV isolates belong to the Asian lineage that was circulating in the Americas [3]. Our isolates were found to be closely related to the CHIKV sequences from the Bahamas 2014 (KY435470), Mexico 2015 CHR1950 (MG921596) and Mexico 2014 InDRE 51 (KP851709), but clustering in a different subclade. According to the terminology of Sahadeo et al, 2017 [2], our isolates grouped in the Asian/American lineage, which is defined by two amino acid substitutions: E2 V368A and 6K L20M. When comparing our isolates with the consensus sequence, 11 point mutations were found in the nsP1, nsP2, nsP3 and capsid genes, presenting more variants than observed in the structural proteins E2 and E1. In nsP1, we observed two synonymous mutations in both isolates (2107 and 2115). nsP2 had one synonymous mutation only in the 2107 sequence. The nsP3 gene had four mutations, three of which were synonymous in the 2107 sequence and one nonsynonymous mutation in isolate 2115. Sequence 2115 mutation is classified as transition and transversion in region 378 of nsP3. There is a change of histidine (hydrophobic amino acid, no polarity) by proline (amino acid hydrophilic with positive polarity), which is a novel finding. Substitution of a hydrophobic residue with a polar amino acid could represent possible structural changes, as observed in other alphaviruses [41], which affects in vitro replication and viral pathogenesis. The observation warrants future studies on CHIKV.

The CHIKV isolates of this study did not group with other isolates from coastal Chiapas which have the E2 V113A substitution as observed for CHIKV isolates in the British Virgin Islands sequences [2]. We also found differences in the non-structural (nsP3 and nsP4) and structural proteins (E2 and E3) of our strains. These findings showed that different strains of the CHIKV Asian lineage were circulating simultaneously in the Central Valley of Chiapas.

In our study, CHIKF cases were diagnosed among people of wide age ranges, from nursling to octagenarians (x̅ = 33 years), supporting the expected outcome for the age distribution of cases due to recent introduction of CHIKV into the completely naïve population of the Americas. A higher proportion of our cases occurred in the age ranges 1-9, 30-39 and 40-49, unlike the data of the Mexican Epidemiological Surveillance System SINAVE/DGC [17], the most affected age group was 25-29 years in 2015. Our findings of clinical manifestations including high fever, polyarthralgia, headache, myalgia, rash, and digestive disorders, matched with the clinical signs/symptoms of CHIKF outbreaks (see table 5 for Confidence 95% intervals) involving the Asian lineage [42–46]. Interestingly, we also observed signs/symptoms that were rarely described in other studies: conjunctivitis of 1-4 days of duration in 62.7% and adenopathy in 41.8%, the later associated with cervical (retroauricular, neck), axillary, and inguinal region. Other studies have reported the development of adenopathies in CHIKF as being uncommon [47,48] whereas such changes are commonly found in other alphaviruses patients, such as ónyong-nyong fever [49]. Also, unlike other studies we found 62.7% of patients with mainly maculopapular and pruritic rash and bullous and pruritic in a schooler patient. One sign that appeared nearly only in women at a 4:1 ratio with men was facial hyperemia. Others were ulcers (17.9%), hemorrhages (6.0%), gum pain (11.9%), burning skin (7.5%) and jaundice (3.0%). The clinical signs of the patient from whom strain 2115 was obtained with the mutation in the nsP3 region were symmetric bilateral arthralgia with severe joint pain in the shoulder, trunk, back and hip; pruritic maculopapular rash in the chest, trunk and abdomen, adenopathy, burning sensation on the skin, high and low back pain, dizziness, nausea, vomiting, difficulty breathing, and lymphopenia. As previously mentioned, clinical signs such as adenopathy, skin burning, and difficulty breathing have rarely been associated with CHIKV infection [11,42,43,45].

In our study, clinical signs and symptoms were observed more frequently in women (60%) than in men, which potentially reflects greater exposure to mosquito bite risk due to the fact of household activities, clothing practices, and/or greater willingness to seek medical care. Also, during the initial phase, we observed that 40% of our CHIKV-positive patients were misdiagnosed by the primary care medical staff. The treating health personnel recorded signs and symptoms due to other etiologies, including DENF, respiratory diseases, stress and chronic exhaustion, common headache, food allergies; and only 15% were properly diagnosed as CHIKF cases. A significant decrease in labor productivity due to absenteeism and disabling pain was also observed, suggesting a serious effect associated with CHIKF at the family household economy.

Misdiagnosis can greatly affect the clinical picture of infected patients. This would impact the evolution from DENF to severe dengue hemorrhagic fever (DHF) disease and the risks unsuitable medication of CHIKF patients with arthralgia-alleviating nonsteroidal anti-inflammatory drugs, which could lead to severe bleeding in patients with DHF or thrombocytopenia [50].

Chikungunya continues to be a latent and important public health problem, especially in regions lacking proper medical care and diagnostic facilities that characterize the Mexican Republic and mosquito vectorial populations that are abundant.

The SINAVE in 2015 reported 683 cases in Chiapas, out of the total number of CHIKF patients registered throughout the country. Between 2016-2018, 18 cases of CHIKF were documented in Chiapas. No cases were recognized between 2019-2021 and until 2022-2023 only two cases, one per year, were reported. As of the epidemiological week 27, 2024, no cases of the CKIKF had been documented [17].

In Mexico, there is an apparent decrease in the circulation and transmission of CHIKV which can be explained mainly by two facts, a) the CDC-PAHO guidelines proposing to test only 5% of the supposedly suspected CHIKF cases in health settings [23] and b) protection considering that the first explosive outbreaks occurred in 2015-2016 that resulted in herd immunity protection by the exposed population. In this context, we assumed that the true burden of CHIK disease in this Mexican region is unknown and that confirmed cases could be under-reported. It is likely that the burden of CHIK disease is underreported because several positive cases were missed by surveillance systems due to clinical similarities with other endemic viral diseases (dengue, Zika, etc.) [17] in the study area.

The current picture reported in other geographical regions of the Americas have revealed explosive outbreaks of CHIKV, particularly in south American countries, suggesting a latent or even progress outbreaks of CHIKV forecasted for Mexico. According to PAHO data from January 1 to September 10, 2024, 11,574,374 cases of CHIKF and more than 6,542 deaths were documented from 20 countries most from the South American region. In 2023, 410,754 including 419 deaths, cases were reported, this was an increase in more than 135,000 cases as compared with the previous 2022 year, with the most affected countries being Brazil, Paraguay, Argentina, Uruguay and Bolivia [17, 51]. Of the five countries, Brazil stands out as one of the most important contributing human migratory numbers into south Mexico, including the Chiapas State region (UN, IOM Annual Report 2024) [18], with trends pointing to in land migratory movements towards the Mexico-US northern border. We believe the picture during 2024 is complemented by an ongoing DENF outbreak near the south Mexican border, including the State of Chiapas, with an increase of 354 % DENF cases only in the July month of 2024 [22,211 DENF cases] as compared to 2023 (4,892 DENF cases) [51]. The actual scenario recorded in the Mexican Republic reinforces the notion that hundreds of cases of CHIKF might be hidden under the dengue umbrella. During our studies in 2015, we found at least one coinfected case of CHIKV-DENV and half of the positive CHIKF cases showed IgG DENV antibodies suggesting that in the 2024 Mexican scenario, DENV diagnosis should be complemented throughout a concerted approach of differential diagnostics with CHIKF by the different regional health entities. We consider this to be an essential approach to help clarify some of the enigmas associated with the explosive arboviral outbreaks going on in many regions of Mexico and elsewhere in the continent. All in all, the scope of CHIKV in Mexico likely represents an underestimation of CHIKF cases. Therefore, it is advisable that differential diagnostics be implemented by the health agencies to distinguish between DENV (flavivirus) and CHIKV (alphavirus) infection. It is important to stress that in the case of any foreseen outbreak of CHIKF in the Chiapas region, an important component of the surveillance policies should focus on molecular approaches to monitor for novel mutations that might increase the vector competence and therefore lead to an increase infection rate among the human vertebrate host in the region. This is going to help determine the true regional CHIKV burden and even other arbovirus etiologies in the Mexican southern regions and elsewhere.

## Supporting information

S1 Table. Chikungunya strains list of isolates

## Data Availability

All data produced in this work are contained in the manuscript.

## Acknowledgements

The authors convey their sincere thanks to Teodora Minguito, María Ángeles Murillo, Lourdes Hernández and Francisca Molero.

## Author Bio

MC. Gonzalez-Perez is a doctoral student at the Universidad Autónoma del Estado de México, where she has been investigating emerging and reemerging zoonotic diseases. Her doctoral studies have an emphasis on management of pathogens of biosafety level 3 and 4.

## Author contributions

Conceptualization: Ana Luz Gonzalez-Perez, Jose Guillermo Estrada-Franco

Design experiments: Ana Luz Gonzalez-Perez, Jose Guillermo Estrada-Franco

Data curation: Ana Luz Gonzalez-Perez

Funding acquisition: Jose Guillermo Estrada-Franco, Scott Weaver, Maria Paz Sanchez-Seco

Investigation: Ana Luz Gonzalez-Perez

Methodology: Ana Luz Gonzalez-Perez, Ana Vazquez, Maria Paz Sanchez-Seco, Scott Weaver

Lab molecular diagnostics: Ana Luz Gonzalez-Perez, Jessica A. Plante, Kenneth S. Plante, Scott Weaver, Douglas Watts, María Paz Sanchez-Seco, Ana Vazquez, Anabel Negredo, Pedro M. Palermo, Jose Guillermo Estrada-Franco

Lab serological diagnosis: Ana Luz Gonzalez-Perez, Fernando de Ory, Jessica A. Plante, Douglas Watts, Jose Guillermo Estrada-Franco

Project administration: Jose Guillermo Estrada-Franco

Resources: Ana Luz Gonzalez-Perez, Scott Weaver, Maria Paz Sanchez-Seco, Douglas Watts, Jose Guillermo Estrada-Franco

Software: Jessica A. Plante, Kenneth S. Plante

Writing – original draft: Ana Luz Gonzalez-Perez, Jose Guillermo Estrada-Franco

Writing – review & editing: Ana Luz Gonzalez-Perez, Douglas Watts, Scott Weaver, Fernando de Ory, Jose Guillermo Estrada-Franco

## Supporting information

S1 Table. Chikungunya strains list of isolates. (XLSX)

